# Genetic Profiling and Early Detection of Type 2 Diabetes Subtypes through Sex-Stratified GWAS and Explainable AI

**DOI:** 10.1101/2025.07.24.25332120

**Authors:** Lorena Alonso-Parrilla, Miguel Ángel Pérez-Elena, Mohammed Yousef Salem Ali, Maedeh Mashhadikhan, Nicolás Gaitán, Leila Satari, Rodrigo Martín, Anthony Piron, Xavier Farré, Natalia Blay, Lydia Ruiz, Aikaterini Lymperidou, Cecilia Salvoro, Rafael de Cid, Josep Lluís Berral, Juan R González, Ignasi Morán, Miriam Cnop, David Torrents

## Abstract

Type 2 diabetes (T2D) is a complex and clinically heterogeneous disease. Although clustering approaches have defined clinical subtypes, their genetic and molecular architectures remain poorly understood. Efforts using genome-wide association studies (GWAS) have been constrained by modest sample sizes and conservative significance thresholds, limiting subgroup resolution. Leveraging recent expansions in cohort scale and diversity, we developed a new analytical framework that integrates GWAS with Machine Learning and eXplainable Artificial Intelligence. Applied to the UK Biobank, this strategy enabled the first comprehensive genetic and molecular characterization of T2D subtypes, identifying 184 genes that define each subgroup’s molecular landscape and associated tissue profiles. In addition, the framework improved detection of T2D risk compared to current Polygenic Risk Scores. The findings underscore the importance of clinical stratification to uncover the complex pathophysiology of T2D and to pave the way for more precise prevention and treatment strategies.

---

Type 2 diabetes (T2D) is a complex metabolic disorder which affects 828 million individuals worldwide ^1^. As a result, T2D is among the top ten causes of death, having caused 3.4 million deaths globally in 2024 ^2^. Hyperglycemia is the key diagnostic marker for T2D, that develops as a consequence of variable degrees of insulin resistance and pancreatic beta-cell dysfunction. T2D is a highly heterogeneous disease, at the molecular and clinical level. This heterogeneity has led to the identification of five clinical subgroups of diabetes: Severe Autoimmune Diabetes (SAID), Severe Insulin-Deficient Diabetes (SIDD), Severe Insulin-Resistant Diabetes (SIRD), Mild Obesity-related Diabetes (MOD), and Mild Age-Related Diabetes (MARD) ^3^. While these clustering approaches have attracted great interest and have been replicated widely, the etiologies of these subtypes of diabetes remain largely unknown. A deeper understanding of the multiple genetic and molecular backgrounds of these subgroups is crucial for improving the management of the disease and to find novel therapeutic targets ^4^. Aligned with this hypothesis, previous studies have suggested that different genetic profiles of the subtypes underlie etiological differences ^3,5–7^.

Despite these advances, exploring the genetic and molecular basis of T2D clinical subgroups at a genome-wide level remains challenging. So far, studies have drawn conclusions from a limited set of known T2D-associated variants, suggesting potential genetic differences between the clinical subgroups ^3^. These genetic differences were also supported by a broader genome-wide association study (GWAS) inspection of subgroups, which reported four previously known T2D associated loci falling in *HLA-DQB1*, *TCF7L2* and *LRMDA* regions with significantly different effects in the subgroups ^6^. Notably, SNP-wide heritability estimates in the same study revealed differences across subgroups, with SIDD and MOD exhibiting higher heritability, while SIRD and MARD lower heritability. The comparison between subgroups of multiple Polygenic Risk Scores (PRS) encompassing T1D, T2D, insulin secretion and sensitivity measures, lipids, weight-related phenotype and diabetic complications ^3,6^ as well as the use of subgroups clustering based on T2D genetic information in larger cohorts ^5^, such as the UK Biobank, has highlighted molecular differences and association with distinct clinical outcomes ^5^. While significant progress has been made in identifying clinical T2D subgroups, their genetic and molecular profiles remain largely uncharacterized.

Methodological limitations of GWAS, including the reliance on highly restrictive *p*-value thresholds (*p*<=5×10^-8^) and other challenges derived from the quality of the data, have left the genetic investigation of T2D clinical subgroups at a genome-wide level largely unexplored. Indeed, relatively small sample sizes of T2D subgroup cohorts limited the power of GWAS to reveal subgroup-specific T2D variants ^6^. This requires the relaxation of *p*-value thresholds to improve discovery rate, which, in turn, increases the false-positive rate. The lack of data on subgroup-defining variables in most cohorts, coupled with their dependence on population-specific characteristics, present a major challenge to identify the subgroups and, consequently, to investigate their genetic and molecular differences ^7^. To overcome these limitations it is essential to explore new analytical frameworks that complement GWAS, expanding the possibilities of marker discovery. Hence, we hypothesize that applying novel methods to large-scale cohorts will fill subgroup-specific gaps by advancing the genomic analysis of T2D clinical subgroups, thereby uncovering their genetic heterogeneity, and will lead to better T2D classification.

The objective of this study is to explore the genetic and molecular landscape of T2D subtypes, overcoming the limitations of traditional GWAS significance thresholds. To this end, we have developed and applied a novel analysis frame based on a combination of mixed logistic regression models (GWAS) with Machine Learning (ML). This combined approach is designed to identify and prioritize biologically relevant genetic and molecular factors contributing and defining the pathophysiology of T2D clinical subgroups. The insights gained from this analysis are expected to advance our understanding of the disease, thereby enabling more accurate risk prediction and refined patient stratification.

## Results

### Genetic characterization of T2D clinical subgroups

As depicted in **Fig. 1A**, to broadly explore the genetic and molecular architecture of T2D subgroups, we first collected genotype array information from 500,000 participants in the UK Biobank ^8^, including 31,344 individuals of European ancestry with diagnosed T2D (**<u>Methods</u>**). We applied genotype array data quality controls ^9^, and imputed high-quality variants ^10,11^ using five reference panels ^12–16^. We next clustered the individuals with diabetes into clinical subgroups using a previously described strategy ^3,17^, which relies on HbA1c levels, body mass index (BMI) and age at onset of T2D ^18^. To enhance subgroup-specific characteristics, we excluded 25% of individuals exhibiting clear characteristics of two or more subgroups (**Supplemental Fig. 1**, **Supplemental Table 1**). We retained a total of 20,755 individuals with non-autoimmune T2D for downstream analyses, including 2,812 SIDD, 6,014 SIRD, 1,689 MOD, and 10,240 MARD cases. Additionally, 233,284 non-diabetic individuals were included as controls. 40% percent of individuals within the subgroups were genetically female, and the distribution across subgroups was comparable between sexes, with a predominance of MARD, followed by SIRD, SIDD and MOD. In line with previous findings ^19^, SIDD and MARD were more prevalent in males, whereas SIRD and MOD showed a higher frequency in females (**Fig. 1B**). The distributions of HbA1c, BMI and age at onset of T2D share overall cluster characteristics with subgroups from clinical cohorts ^3^.

**Figure 1.**
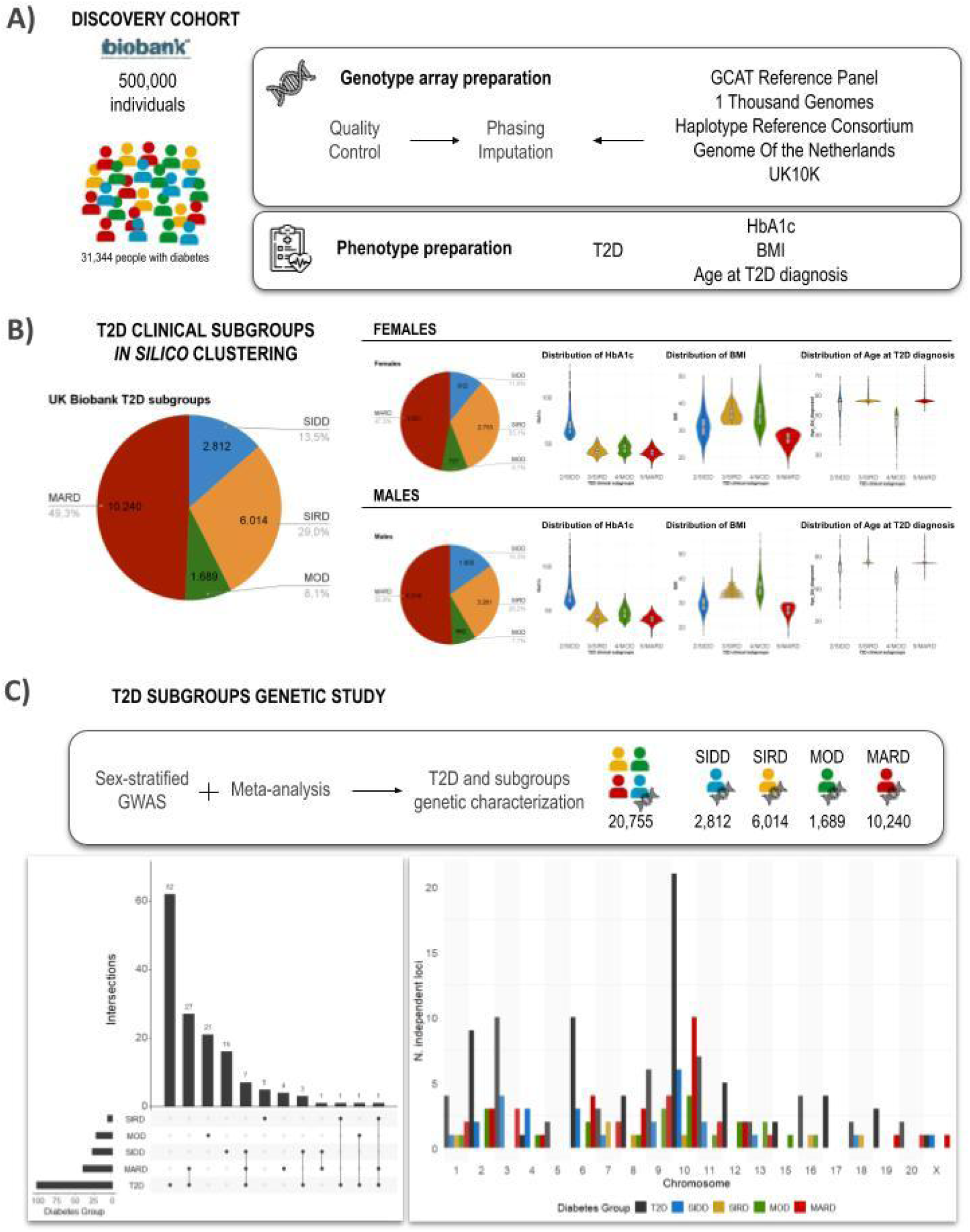
Overall GWAS strategy for T2D subgroups. **A)** We used genetic and clinical information from participants in the UK Biobank (500,000 European ancestry individuals; 31,344 with T2D). **B)** We used HbA1c, BMI and age at onset of T2D to classify diabetic individuals into clinical subgroups. Pie charts display the proportion of each T2D cluster within the UK Biobank. Violin plots show the distribution of clinical variables (HbA1c, BMI, age at onset of T2D) for each subgroup for males and females. **C)** We characterized the genetics of T2D subgroups by testing their genomic association with disease through sex-stratified GWAS followed by meta-analysis. The UpSet plot on the left displays the intersection between independent loci found in T2D and distinct clinical subgroups. The barplot on the right illustrates the number of independent loci found in T2D and clinical subgroups by chromosome.

Using Firth logistic regression sex-stratified analyses ^20^, followed by meta-analysis ^21,22^ for a sex-combined approach (**<u>Methods</u>**), we tested 15,586,493 high-quality genetic variants for association with overall T2D as well as with each of the four clinical subgroups separately. We found a total of 149 significant (*p*<=5×10^-8^) independent loci, of which 102 significant in the full cohort, and 27, 7, 22, and 40 loci in the SIDD, SIRD, MOD, and MARD subgroups, respectively. Of these, 40 loci were shared between the overall cohort and at least one subgroup, while 62 were exclusively detected in the full cohort and 47 in subgroups (**Fig. 1C**, **Table 1**, **Supplemental Fig. 2-6**). From all significant loci identified in this study, 88 were previously reported significant in T2D analyses ^6,9,23–26^, while 61 represent novel discoveries.

### Genetic heterogeneity of T2D subgroups

Beyond enabling the identification of novel T2D-associated loci masked by analyses of the overall cohort, the subgroup-specific approach also revealed substantial genetic and molecular heterogeneity among the distinct disease subtypes. From all 87 loci identified in at least one subgroup, 9 were shared between at least two of them, whereas 19, 6, 22, and 31 loci were identified only in the SIDD, SIRD, MOD, and MARD subgroups, respectively (**Table 1**, **<u>Methods</u>**). SIDD and MARD exhibited a greater degree of genetic overlap, whereas SIRD and MOD appeared to be more genetically distinct (**Fig. 2A**). Even at an exploratory significance threshold (*p*<=5×10^-4^), only 33 loci were shared among at least two of the clinical subtypes, of which 27 (82%) were shared between SIDD and MARD.

**Figure 2.**
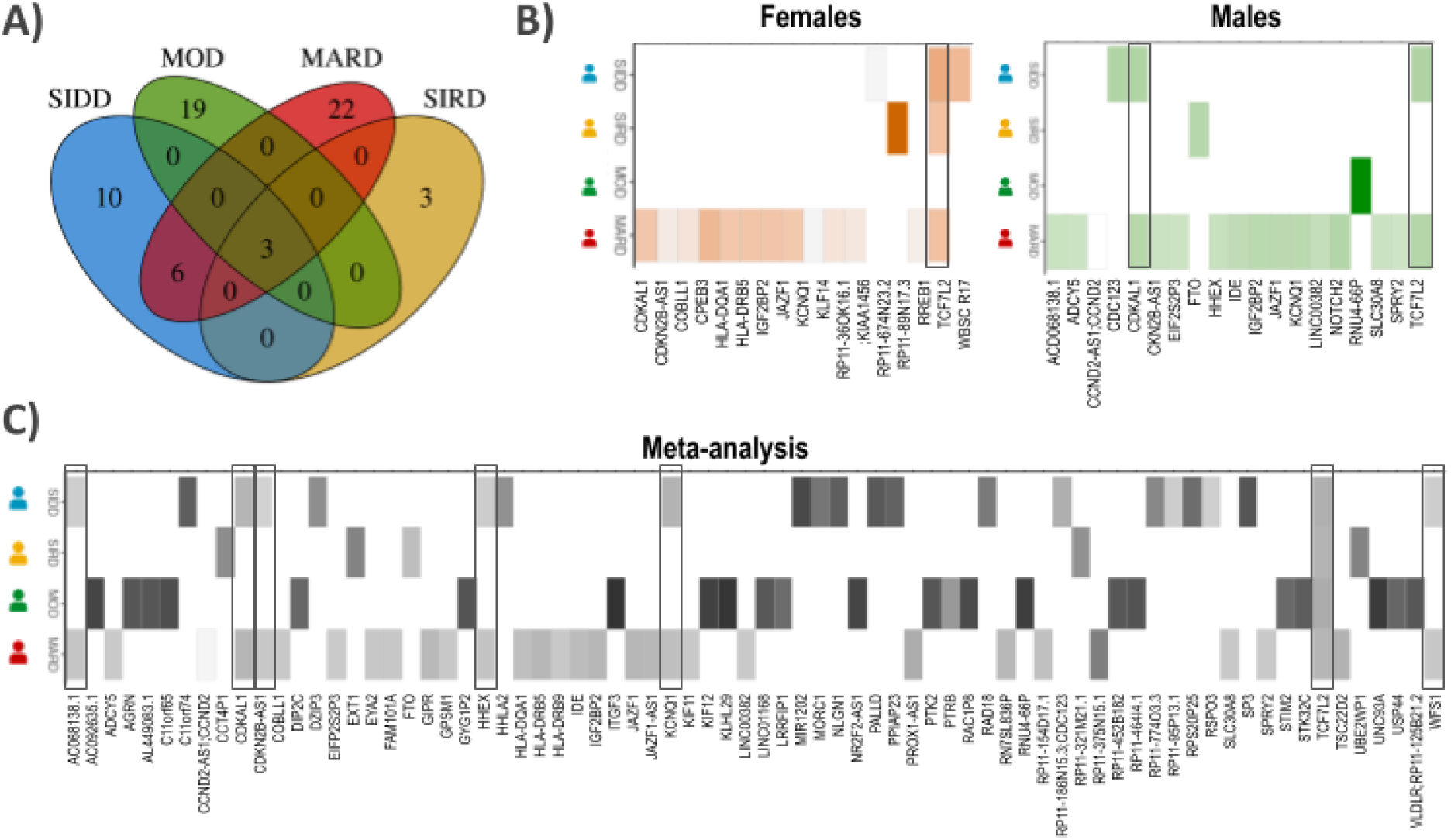
Genetic and molecular heterogeneity of T2D subgroups. We annotated the genes linked with significant variants using annotations from the Translational Human Pancreatic Islets Genotype Tissue Expression Resource (TIGER), Genotype-Tissue Expression Project (GTEx) and Ensembl database. **A)** Venn diagram illustrates the loci intersections between clusters. Heatmaps display the average effect in significant variants from each specific gene region for each subgroup in **B)** sex-stratified GWAS (orange females and green males), and **C)** meta-analysis (gray). A high intensity indicates more risk of disease. Squared regions highlight shared loci across subgroups.

We further examined whether the nine loci shared across the diabetes subtypes contribute to their molecular heterogeneity by comparing their effect sizes across subgroups **Fig. 2B-C**. Three loci, located within an intronic region of *TCF7L2*, were common to all clusters. The remaining six loci were significant only in SIDD and MARD; five of these were in strong linkage disequilibrium (LD) (R^2^>=0.8) with expression quantitative trait loci (eQTLs) for the genes *AC068138.1*, *WFS1*, *CDKAL1*, *CDKN2B-AS1*, and *HHEX*. The sixth locus was located in proximity to *KCNQ1* (**<u>Methods</u>**). Although all six loci exhibited consistent effect directions, their association magnitudes differed between SIDD and MARD, underscoring subgroup-specific variation (**Table 2**, **Supplemental Table 2**). Variants within the *TCF7L2* region showed higher median effect sizes in SIDD and MOD compared to SIRD and MARD (𝑚(β_𝑆𝐼𝐷𝐷_)= 0. 19, 𝑚(β_𝑆𝐼𝐷𝐷_) = 0. 14, 𝑚(β_𝑆𝐼𝐷𝐷_) = 0. 25, 𝑚(β_𝑆𝐼𝐷𝐷_)= 0. 12). Stratification by sex revealed further subgroup-specific differences: in females, *TCF7L2* variants were associated with SIDD, SIRD, and MARD, whereas in males, associations were limited to SIDD and MARD. Consistent with prior findings linking *TCF7L2* ^27^ to insulin resistance and its sex-specific role in glucose homeostasis ^28^, we observed sex-differentiated heterogeneity (*p*<0.05) for one of the lead SIRD variants, rs61875120, which showed a stronger effect in females (OR=1.20, *p*=1×10^-9^) than in males (OR=1.10, *p*=1×10^-3^).

### Machine Learning enhances disease genetic exploration

To expand upon GWAS findings, we refined current analytical approaches to identify genetic candidates that met nominal significance thresholds but not genome-wide significance (*p*<=5×10^-8^). While these regions have the potential to include novel and unforeseen loci, they carry an elevated risk of false positives, necessitating stringent filtering and validation to ensure biological relevance. To address this challenge, we integrated GWAS data with a ML framework.

To characterize genetic contributors to T2D and each T2D subgroup, we selected an exploratory set of variants from the meta-analysis (*p<=*5×10^-4^) and trained XGBoost ^29^ to classify individuals as diseased and non-diseased (**<u>Methods</u>**). This interpretable mixed-model approach prioritized variants based on predictive performance rather than GWAS p-values alone, enabling the identification of subgroup-associated loci and corresponding classification scores (**Fig. 3A**). To ensure robustness, we replicated and cross-cohort validated the classifier in T2D (**Supplemental Fig. 7**, **<u>Methods</u>**). Overall, our analysis indicated that this method prioritizes variant selection based on predictive performance, which is expected to correlate with effect sizes and variants combination, rather than relying solely on GWAS *p*-values (**Supplemental Fig. 8-9**).

**Figure 3.**
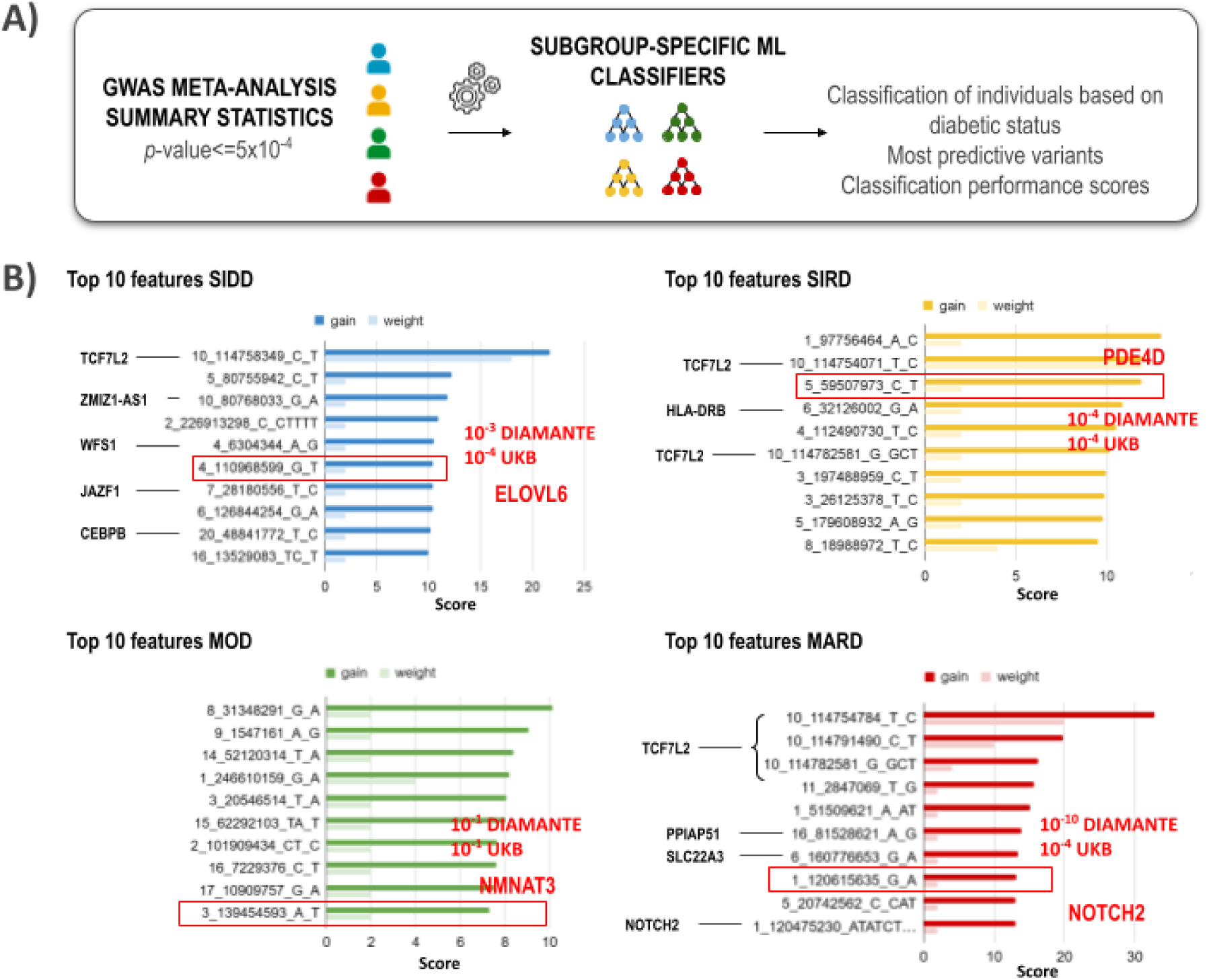
Method interpretability and genomic variant contribution to T2D. **A)** To broaden the genetic characterization of T2D subtypes, we selected the most predictive variants from the meta-analysis (*p<=*5×10^-4^), and used a ML model approach to generate subgroup-specific classifiers. After this analysis, for each subgroup we obtained the list of the most relevant variants to classify the diabetes subtype, a classification of individuals in diseased and non-diseased and the corresponding classification performance scores. **B)** Top ten features obtained from the ML model trained in each clinical subgroup. Bar plots represent the gain (dark) and weight (light) scores assigned by the model to each genomic position (hg19). We only annotated genes linked with GWAS significant variants in UK Biobank (*p*<=5×10^-8^). We used annotations from the Translational Human Pancreatic Islets Genotype Tissue Expression Resource and the University of California Santa Cruz genome browser.

Applying this strategy to the subgroups, we identified 1,359 loci contributing to SIDD (68% sensitivity), 1,477 loci to SIRD (61% sensitivity), 1,132 loci to MOD (68% sensitivity), and 1,565 loci to MARD (63% sensitivity) (**Supplemental Table 3**). Notably, only 9% of these prioritized loci reached the genome-wide significance in our analysis or in previous large-scale T2D GWAS meta-analyses ^9,23,24,26^ (**Fig. 3B**). For example, rs12108641 was among the top ten variants detected by the ML model in SIDD, but not reach genome-wide significant association in our analysis (sex-combined OR=1.18, *p*=1.4×10^-4^), nor in the DIAMANTE study ^26^ (OR=1.10, *p*=3.9×10^-3^). Of note, this variant falls in the 3 prime UTR region of *ELOVL6*, a fatty acid elongase that plays a crucial role in energy metabolism and insulin sensitivity, supporting its role as a fundamental marker for this subtype ^30,31^.

Remarkably, although many of the loci identified by our model had not previously reached genome-wide significance, the full set of prioritized variants was significantly enriched (*p*<2.2×10^-5^) in pancreatic islet regulatory regions and eQTLs, suggesting a biological relevance to T2D. Specifically, 37% of the prioritized loci were in strong LD (R^2^>=0.8) with an already known eQTL (5% FDR; 2,019 loci) and more than 20% of the variants were falling in islet regulatory regions ^11,32–34^ (**Supplemental Fig. 10**).

### Integrative GWAS and ML reveal the molecular background of T2D subgroups

To characterize the functional and molecular landscape of T2D subgroups, we harnessed the interpretability of our integrative methodology to systematically interrogate the biological pathways associated with the loci prioritized by our ML approach, alongside those identified through GWAS significance. By integrating data from 49 tissues in GTEx ^34^ and pancreatic islets from TIGER ^11^, we linked genetic variants to their respective eQTL genes (R^2^>=0.8) and analyzed their functional enrichment (**<u>Methods</u>**). We observed that our ML framework facilitated a more refined characterization of the molecular landscape across subgroups, revealing additional subgroup-specific genes and enriched pathways that remained undetected in standard GWAS analyses, even when analysing only marginally significant loci (*p*<=5×10^-4^). These findings include subgroup-specific colocalizing eQTLs, such as *GCKR*, *MADD*, *SNX19* and *APOE*. In general, we observed enrichment (*p*<=0.01) of several biological processes, including functions that align with the known nature of each clinical subgroup (**Fig. 4**, **Supplemental Table 4**, **Supplemental Fig. 11-14**). For example, we found a significant enrichment of pathways involved in glucose metabolism and insulin secretion in SIDD, consistent with the crucial role of pancreatic islet beta-cell dysfunction in this subgroup. Reflecting the well-established link between insulin resistance and altered lipoprotein dynamics, genes associated with SIRD were predominantly enriched in HDL metabolism pathways. In line with the metabolic profile of MOD, genes associated with this subgroup showed significant enrichment in pathways related to adiposity, obesity, and lipid metabolism. MARD genes showed an enrichment in functions implicated in age-related disorders, cell cycle regulation, and catabolic processes, highlighting their potential role in aging-related metabolic dysfunctions.

**Figure 4.**
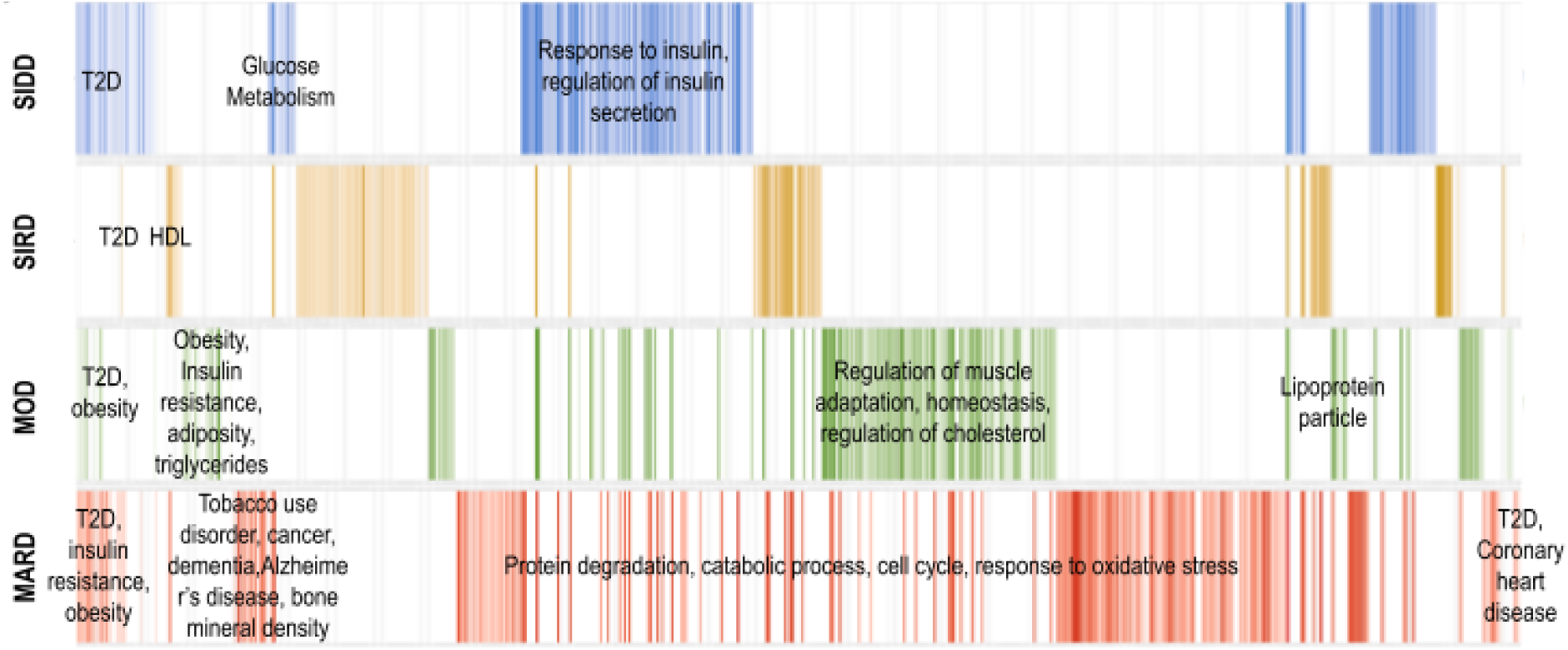
T2D subgroup molecular heterogeneity. Heatmaps represent gene counts for significantly enriched functions in each subgroup (*p*<0.05). MARD enrichment is represented in red, MOD in green, SIRD in yellow and SIDD in blue. The color intensity represents the number of genes supporting the functional enrichment. A high intensity indicates strong functional enrichment.

Beyond the molecular patterns outlined above, this functional enrichment analysis provides a comprehensive catalog of genes and associated functions across T2D subgroups in **Supplemental Tables 4-5**. These datasets offer novel insights into T2D genetics and heterogeneity, which might implicate novel biological functions. For example, the SIDD subgroup shows specific pathway enrichment in folate transport and metabolism, driven by genes such as *DHFR2*, which has been recently implicated in allelic expression imbalance under both acute and chronic hyperglycemic conditions ^35^. More broadly, the functional landscape across T2D subtypes reveals both shared and distinct enriched biological processes. As could be expected, the general term for energy-related metabolic pathways is common among SIDD, SIRD, and MARD (e.g., R-HSA-1480728). In contrast, unique enriched terms include extracellular matrix remodeling (e.g., collagen and anchoring fibrils) in SIDD, cell cycle progression and senescence-associated signaling (e.g., ORC complex assembly and SASP) in SIRD, cytoskeletal regulation, immune activation, and vesicle transport (e.g., Rho GTPase signaling) in MOD, and insulin and oocyte signaling (e.g., progesterone-mediated oocyte maturation) in MARD.

To further investigate the tissue-specific function of variants in subgroups, we performed colocalization analyses between the T2D subgroups loci and eQTLs in 50 tissues from GTEx and TIGER ^11,34^. This identified 184 genes linked with diabetes subtypes (PP.H4>=0.8, **Fig. 5A-B**, **Supplemental Table 5**, **Supplemental Fig. 15-18**, **<u>Methods</u>**), offering insights into potential subgroup-specific mechanisms (**Fig. 5C**). The limited overlap of colocalizations in tissues underscores the unique molecular makeup of the subgroups. Genes associated with SIDD predominantly colocalized within pancreatic islets, with over 20% of genes mapping to this tissue, while 7% in MOD, 13% in MARD and only 2% for SIRD, keeping with the central role of beta-cell dysfunction in SIDD, its contribution to MOD and MARD and its limited involvement in SIRD. SIRD genes showed 28% colocalization within skeletal muscle, a tissue central for insulin sensitivity. MOD-associated genes displayed strong colocalization in tissues implicated in vascular dysfunction and obesity-related complications, including visceral adipose tissue (28%), that is central to metabolic impairments in obesity and T2D, and 21% in vascular tissues (aorta and tibial artery), while MARD genes were mapped to thyroid, nerve and adipose tissue, with over 20% of genes colocalizing these tissues.

**Figure 5.**
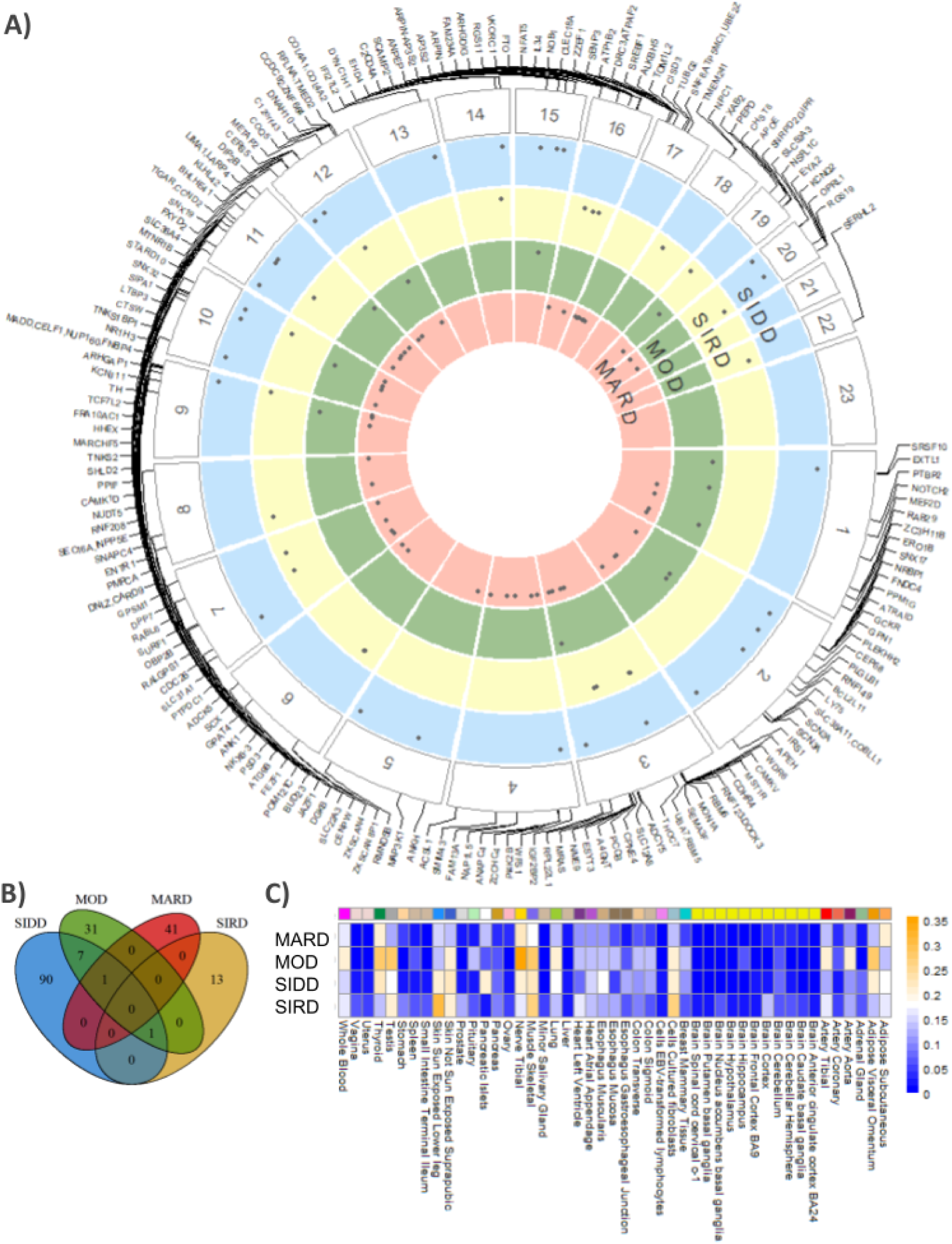
T2D subgroup colocalizing genes. **A)** Circle plot of colocRedRibbon colocalizations of GTEx and TIGER eQTLs with T2D GWAS combined ML strategy. eQTL genes are positioned at their starting position on the chromosome. The concentric circles correspond to each subgroup (SIDD in blue, SIRD in yellow, MOD in green and MARD in red). Each dot corresponds to a SNP. **B)** Venn diagram with the overlap between T2D subgroup colocalizing genes. **C)** Heatmap represents the proportion of colocalizing variants in tissues for each subgroup. The color scale represents the proportion of colocalizing genes in that tissue for each specific subgroup, with blue for values below and orange for values above the average.

**Figure 6.**
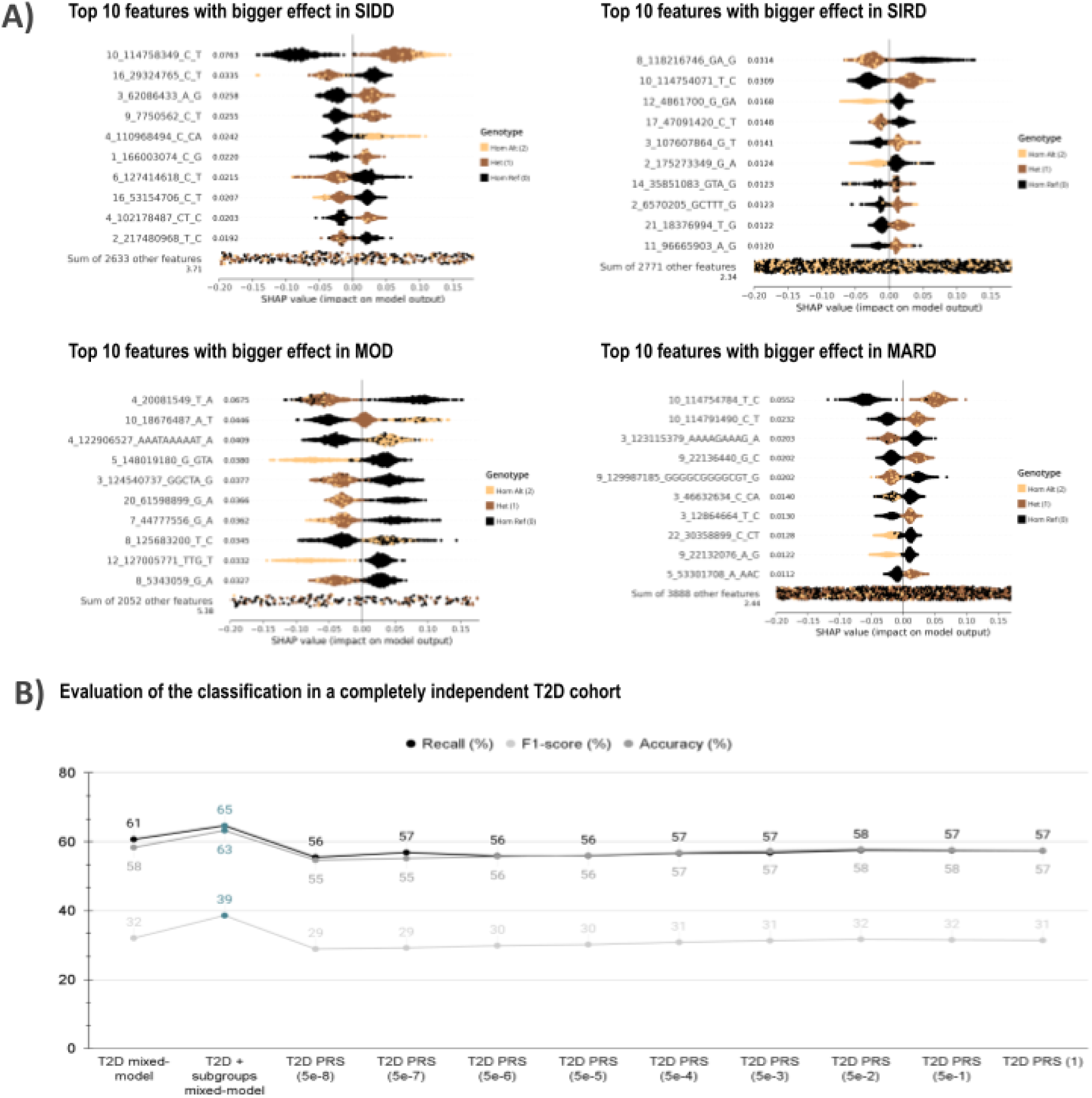
T2D machine learning prediction. **A)** Top ten variants with higher contribution to individual classification. Each dot represents one individual and the contribution of the genotype, being homozygous reference (black), heterozygous (brown) and homozygous alternate (yellow), to the final classification. A negative SHAP value represents that the genomic position (hg19) contributes negatively to the diabetic classification and, therefore, reduces disease risk, while a positive SHAP value indicates that the variant increases disease risk. The value next to each genomic position (hg19) represents the median of the absolute value of the variant contribution to the classification. **B)** Scatterplot represents the comparison of classification results obtained between different predictors trained in UK Biobank and tested in 70KforT2D. Results obtained from the combination of the general T2D mixed-model with the subgroup predictors are highlighted in blue.

### Impact of subgroup-specific information in T2D prediction

Prediction of T2D before symptoms onset, and specifically of the T2D subgroup type, is of utmost clinical importance for disease outcome and treatment. For this reason, we sought to evaluate subgroup-specific prediction of disease. To this end, we employed the pre-trained models for each T2D subgroup to perform predictions on subgroup-specific independent test sets derived from the UK Biobank. Compared to current T2D PRS, the subtype predictors increased sensitivity by 6%, achieving sensitivities of 68%, 61%, 68% and 63% for SIDD, SIRD, MOD, and MARD, respectively (**Supplemental Table 6**, **<u>Methods</u>**).

To further dissect the genetic architecture underlying these predictions, we next applied eXplainable Artificial Intelligence methods ^36^ (**Fig. 6A**). Through this analysis, we gained insight into the classification criteria of the model, allowing us to determine why an individual was categorized as a case or a control while also quantifying the contribution of each predictor to the decision-making process. For example, we observed that the novel T2D variant nearby *ELOVL6* (rs12108641) confers risk for disease development in SIDD, but not for the other diabetes subtypes. In contrast, variants within the intronic region of *TCF7L2* were consistently informative for classification of SIDD and MARD.

Finally, we aimed to develop a comprehensive model for global T2D prediction. To accomplish this, we classified each individual using both subgroup-specific models and a general T2D classifier, evaluating classification performance on an independent test set that included individuals not involved in any prior training, covering all subtypes (SIDD, SIRD, MOD and MARD). To further improve predictive accuracy, we integrated the outcomes of all these models. Compared to PRS, our integrative model resulted in a 7% increase in diabetes detection and a 5% improvement in classification accuracy in the 70KforT2D dataset (**Fig. 6B**, **Supplemental Table 6**, **<u>Methods</u>**). This highlights once again the value of our integrative approach in expanding the genetic knowledge of T2D and its subgroups.

## Discussion

In this study, we provide the first comprehensive characterization of the genetic and molecular architecture underlying T2D clinical subtypes by applying an integrated GWAS and ML framework to analyze UK Biobank data. This novel approach leverages the strength of ML to recover and interpret hundreds of biologically relevant GWAS signals, including variants that fall below conventional genome-wide significance thresholds and would otherwise be overlooked. Our findings also demonstrate the added value of dissecting T2D into clinically defined subgroups, which substantially increases discovery power by overcoming the dilution of subgroup-specific genetic signals expected to occur in analyses of heterogeneous, mixed T2D cohorts. This strategy enabled the identification of both shared and subgroup-specific variants, providing distinct genetic and molecular profiles for each clinical subtype.

Our results reveal considerable heterogeneity in the genetic architecture of T2D subtypes. Even among variants shared across subgroups, we observe substantial differences in effect sizes, underscoring the molecular complexity and diversity underlying these clinically distinct forms of T2D. Beyond confirming established molecular pathways associated with each subgroup, we identify novel signals that may point to previously unrecognized biological mechanisms and potential therapeutic targets, like extracellular matrix remodeling ^37^ and folate transport in SIDD through *DHFR2,* recently related with allelic expression imbalance under hyperglycemic conditions ^35^; senescence-related signaling in SIRD, immune and cytoskeletal regulation in MOD, and reproductive hormone signaling in MARD.

Our tissue-specific analyses further support the distinct molecular signatures characterizing each subgroup, with subgroup-specific enrichment observed in pancreatic islets, skeletal muscle, thyroid, nerve, adipose tissue, aorta and tibial artery. Variability in tissue-specific function aligns with previous studies demonstrating systemic transcriptomic alterations in T2D ^38^. Importantly, the subgroup-specific differences in tissue expression profiles provide insight into potential subgroup-dependent biological processes and complications, reinforcing the value of stratified analyses for understanding disease mechanisms.

Additionally, our ML approach allowed integrating multiple loci in risk prediction models, which improved the identification of individuals at elevated T2D risk by 7%. It is important to note that these risk models were constrained to variants surpassing a predefined association threshold (*p*<=5×10⁻⁴), which may have excluded relevant genetic signals. Future studies applying this approach at a genome-wide scale are warranted to uncover novel biomarkers and further refine risk prediction models.

This study has several limitations that should be acknowledged. First, even though we selected only individuals with well-defined, non-overlapping subgroup characteristics, the absence of key metabolic parameters, such as insulin secretion and sensitivity (HOMA-IR and HOMA-B) and glutamic acid decarboxylase autoantibodies, may have reduced the resolution of our clinical stratification. Second, our analyses were limited to common genotyped variants, and the potential contribution of rare or structural variants to subgroup heterogeneity cannot be excluded. A third limitation is the focus on individuals of European ancestry, which limits the extrapolation of our findings to other populations, where both shared and ancestry-specific genetic effects are likely to exist. Finally, statistical power, particularly within smaller subgroups, remains a limiting factor. Future studies with larger, multi-ethnic cohorts and full clinical data will be critical to validate these results and resolve the genetic and molecular architecture of T2D subtypes. Despite these limitations, the analytical framework in this study enabled the characterization of the genetic and molecular architectures underlying T2D subgroups, and represents a significant advance toward improved risk prediction and precision patient stratification.

## Methods

### UK Biobank cohort

UK Biobank is a large-scale biomedical database and research resource with 500,000 participants, most of them of European ancestry ^8^. This study was performed based on the information from the last update of the cohort in 2023, therefore, this population-based cohort study comprises individuals aged between 50-90 years which were recruited in diverse regions of the United Kingdom in 2006 and 2010. Participants allowed the collection of biological samples, including blood, urine, and saliva, for genetic, metabolomic and proteomic analyses. Participant data include comprehensive information on their health, lifestyle and environment, and health records. All study participants provided informed consent. Individuals with Type 2 Diabetes were selected using the International Statistical Classification of Diseases and Related Health Problems 10th Revision (ICD-10), category E11.

### 70KforT2D cohort

The 70KforT2D is a T2D case-control dataset which includes data from 12,926 diabetic and 57,191 non-diabetic individuals of European ancestry ^9^. The individuals included in this dataset belong to five studies: Resource for Genetic Epidemiology Research on Aging (GERA), Finland-United States Investigation of NIDDM Genetics (FUSION), Wellcome Trust Case Control Consortium (WTCCC), Gene Environment Association Studies initiative (GENEVA), Northwestern University NUgene project (NUgene) ^39–43^. The genetic information is publicly available through the dbGaP platform for FUSION (phs000867.v1.p1), GENEVA (phs000091.v2.p1), NUgene (phs000237.v1.p1), GERA (phs000788.v2.p3), and the Sanger platform for WTCCC. The available metadata for each individual corresponds to measures of body-mass index (bmi), sex, age and diabetic type. Nonetheless, there is no available information from NUgene individuals’ age, neither for WTCCC individuals’ age and bmi. To avoid the possible overlap of samples between the UK Biobank and 70KforT2D, we remove the individuals from WTCCC cohort, thus accounting with the information of 65,316 individuals (11,037 diabetic and 54,279 non diabetic).

### GCAT|Genomes for life cohort

The GCAT|Genomes for life cohort is a prospective dataset which includes data from 20,000 individuals of European ancestry between 40 and 65 years at recruitment in 2014-2017 ^44^. Participants allowed the collection of blood plasma, serum and white blood cells samples, as well as other clinical and anthropometric measures and to link with electronic health registers from the Catalan public healthcare system. The dataset utilized in the analyses comprises the genotype of 4,988 individuals, among which 450 are diagnosed with diabetes and 4,538 are non-diabetic.

### Genotype Quality Control and Imputation

To ensure the quality of the genotype array data, we followed the strategy in Bonàs-Guarch *et al.,* 2018 ^9^ and used PLINK ^45^ to perform quality control at sample and variant level in the UK Biobank, the 70KforT2D and the GCAT|Genomes for life cohorts.

At the individual level, we discarded individuals with over 5% of non-genotyped variants from their arrays or reporting gender-sex inconsistencies. Inclusion criteria for the study are that a participant must be of European ancestry according to principal component analysis. In the case of the UK Biobank, we only kept individuals, which according to the Pan-UKB study, were from European ancestry ^46^.

At the level of the variant, we excluded variants with a missingness frequency above 5%, and only kept the ones with a minor allele frequency (MAF) higher than 5% and in Hardy-Weinberg equilibrium (HWE) with a Bonferroni corrected *p*-value lower than 10^-20^.

Finally, following the strategy in Pérez-Elena *et al.* (manuscript in progress) we performed an X-specific quality control in the non-pseudoautosomal region. We split the chromosome X genotype by sex and applied the filters previously explained. The HWE filter was only performed on females, and the variants excluded after this filter were also excluded in the males cohort. Finally, variants presenting differences in allele frequency and genotype missingness between males and females were eliminated (*p*<1×10^-6^).

After the quality control, the final dataset from the genotype array consists of 422,090 European individuals with 583,101 high-quality genotyped variants in the UK Biobank, 70,127 individuals in the 70KforT2D and 4,988 individuals in the GCAT|Genomes for life.

After quality control, we used GUIDANCE ^10^, relying on Shapeit4 ^47^ and Impute2 ^48^ to phase and impute the genotype array information. We performed imputation with five different reference panels: GCAT, 1000G phase3, HRC, GoNL and UK10K ^12–16^. We only kept the variants with high quality (INFO>0.7). The results obtained from the different panels were merged following the strategy in Alonso *et al.,* 2021 ^11^, thus, keeping the variants with the best info score obtained across all panels, and filtering the variants with MAF>0.1%.

After this step 15,586,493 variants were kept for downstream analysis in the UK Biobank and 15,131,345 in the 70KforT2D.

### Association test and meta-analysis

To increase the power of detection of sex-specific signals, we performed a sex-stratified GWAS followed by meta-analysis.

For the GWAS analysis, we selected diabetic individuals based on ICD-10 codes, thus, E11 (non-insulin-dependent diabetes mellitus). Then, some exclusion criteria were applied to the cases and controls:

i. to avoid contamination from other types of diabetes mellitus, we only included the non-insulin dependent diabetes mellitus individuals without presenting other types of diabetes E10 (insulin-dependent diabetes mellitus), E13 (other specified diabetes mellitus), and E14 (unspecified diabetes mellitus), and
ii. we only kept as control subjects those individuals with no documented disorders classified under Chapter IV (Endocrine, nutritional and metabolic diseases) and no reported family history of diabetes mellitus, based on UK Biobank data fields 20107 and 20110.

As a result, we kept 143,919 females (131,465 controls and 12,454 with diabetes) and 120,710 males (101,820 controls and 18,890 with diagnosed T2D).

To define the groups of diabetic individuals we applied the clustering methodology presented in Ahlqvist *et al.,* 2018 ^3^. However, since there are no measures available in the UK Biobank for glutamic acid decarboxylase antibodies and HOMA, we generated the clusters based on HbA1c level, BMI at baseline and age at onset of T2D which have been successful in classifying T2D patients in previous studies ^49,50^ (**Fig. 1A-B**). As stated in Ahlqvist *et al.,* 2018 ^3^, to ensure the clinical usefulness of the clusters, we assigned patients to clusters according to ANDIS cluster centroids coordinates.

To that end, the Euclidean distance from the nearest cluster center was used to assign the diabetic patients from the UK Biobank to each cluster. Following the strategy in Kahkoshka *et al.,* 2020 ^50^ to confirm the reliability of our clusters we calculated the subject-specific ratio between the smallest and the second smallest Euclidean distance to the ANDIS cluster centers (**Supplemental Fig. 1**). We only kept individuals with a ratio below 0.8 to ensure their unique participant-cluster-association (**Supplemental Table 1**).

To increase the detection power of genomic variants associated with subgroups, we excluded from the control group individuals with similar traits to the cases, thus, sharing the same chapter, we limited the case-control ratio to 1:100 ^51^ and selected the oldest controls. With this ratio, there would be >80% power to detect common variants (MAF>0.05) with at least 𝑂𝑅ɛ[0. 3, 2. 2]^52^, independently of the prevalence of the disease in the general population. After applying these filters, we kept 233,284 controls (131,465 females and 101,819 males).

We used REGENIE ^20^ for association testing, which allowed the analysis for case-control unbalanced data and can account for sample relatedness and population structure. With the aid of this tool we performed a Firth logistic regression for each sex where each test was adjusted for 7 genetic ancestry principal components calculated with PLINK ^45^.

To increase the statistical power through the combination of male and female samples and allow the discovery of loci that contribute to complex human traits in the population, we used GWAMA (Genome-Wide Association Meta-Analysis) software ^21,22^. This allowed us to estimate male- and female-specific allelic effects based on fixed-effects by estimating chi-square distribution with one degree of freedom.

After the association test and meta-analysis, we obtained a list of variants passing the Bonferroni corrected *p*-value threshold (5×10^-8^). To define each locus, we used PLINK tool ^45^ to perform a variant pruning based on LD and keep only the loci in linkage equilibrium with each other, within a 500Kb window (--indep-pairwise 500 100 0.2).

To distinguish between known or new associated regions, for each significant variant we looked for a proxy variant with an LD R^2^>=0.35, in different datasets. More specifically, we contrasted the novelty of T2D clinical results with Ahlqvist *et al.,* 2018 ^3^ and Aly *et al.,* 2021 ^6^ significant signals. To compare with already known T2D signals, we analyzed the overlap between our results with the latest large T2D GWAS meta-analyses, thus, including the 70KforT2D, DIAMANTE, T2D clinical subgroups and all the T2D GWAS reported signals in the GWAS Catalog (accession 7 June 2024) ^3,6,9,23,24,26^.

To detect heterogeneity in allelic effects between T2D clinical subgroups we used GWAMA^22^. In particular, we used Cochran’s Q-statistic test and I2. We considered as highly heterogeneous the signals with i2>=0.75 (q *p*-value<0.05). The highly sex-differentiated heterogeneous signals were the ones with sex-heterogeneity *p*-value<0.05.

### Gene annotation and functional interpretation

To annotate the variants and, therefore, find and designate locations based on their relations with genes, we prioritized the annotations using diverse resources:

i. Direct relations between the variants and genes relying on their effect in gene expression across different tissues. We used eQTL summary statistics from the Translation human pancreatic Islets Genotype-tissue Expression Resource (TIGER) ^11^ and the Genotype-Tissue Expression resource (GTEx version 8) ^34^. All variants that were in strong LD (R^2^>=0.8) with and eQTL were annotated with the corresponding gene,
ii. Functional impact on genes. We used Variant Effect Predictor (VeP) database ^53^,
iii. Positionally. We used the GWAS Catalog ^24^ and the Ensembl database (GRCh37.p13) ^54^, annotating the variants with the closest gene.

To link the variants with their genetic context, we annotated the outcomes from the analyses relying on their overlap with pancreatic islet regulome annotations and epigenetic marks ^32,33^.

To analyze the functional impact on T2D of the outcomes from GWAS sex-stratified analysis and meta-analyses in clinical subgroups we studied the significant enrichment in T2D transcription factors, biological processes, pathways, regulatory motifs, disease and protein complexes. To that end, we used functional enrichment tools g:Profiler ^55^ and DAVID ^56^. We provided the list of eQTL genes ^11,34^ in strong LD (R^2^>=0.8) with significant variants in each clinical subgroup, excluding the HLA region (hg19 chr6:29691116-33054976) ^57^. In the case of the ML model, we used as the background the GWAS signals provided as input (*p*<=5×10^-4^).

To explore the overlap between the outcomes of our analyses with already known T2D functional genes, we performed colocalization analysis relying on colocRedRibbon ^58^. To obtain summary statistics from the annotated genes, we used GTEx v8 ^34^ and TIGER ^11^, and then performed colocalization analysis with the outcomes from the meta-analysis and sex-stratified GWAS for each subgroup (**Supplemental Table 5**). Only genes with a posterior probability greater than 0.8 were considered for downstream analyses.

### Machine Learning Model

We evaluated the performance of different ML classifiers available from the scikit-learn library in python ^59^ to explore their classification power in T2D and T2D clinical subgroups. These methods included Nearest Neighbors, Linear Support Vector Machine, Radial Basis Function Support Vector Machine, Gaussian Process, Decision Trees, Random Forest, Neural Networks, AdaBoost, Naive Bayes, Quadratic Discriminant Analysis and XGBoost. The best performance in terms of computing time, precision and the data type accepted by the method was obtained by XGBoost ^29^, an explainable supervised ML classifier which relies on random forests.

To prevent errors derived from extremely imbalanced data, we paired diabetic and non-diabetic individuals. Additionally, to overcome current methodological limitations of ML models derived from the high dimension of the variables in contrast with the observations, we only kept variants presenting a certain degree of association with T2D. Therefore, the input of the ML model to find groups of variants associated with T2D consisted of the genotype of 19,014 individuals (9,507 diabetic, 9,507 non-diabetic) in 105,896 variants (*p<=*5×10^-3^) from the 70KforT2D, and the genotype of 66,405 variants (*p<=*5×10^-4^) in55,438 non-related individuals (27,719 diabetic, 27,719 non-diabetic) of European ancestry from the UK Biobank. In the case of T2D clinical subgroups we only worked with the UK Biobank information and retained 7,700 individuals (3,850 diabetic and 3,850 non-diabetic) and 17,383 variants for SIDD, 17,210 individuals (8,605 diabetic and 8,605 non-diabetic) and 16,669 variants in SIRD, 6,236 individuals (3,118 diabetics and 3,118 non-diabetics) and 16,439 variants in MOD and 24,292 individuals (12,146 diabetic and 12,146 non-diabetic) and 27,528 variants in MARD.

A basic train-test algorithm was prepared first splitting the discovery dataset in two independent datasets: a train set and a test set. Two different split values were tested 0.2 and 0.3, keeping 20% and 30% of the data for testing, respectively. The train set was used by the XGBoost algorithm to learn and the test set to evaluate the results. To prevent overfitting of the model and to obtain the best performance, we used a grid search hyper-parameter adjustment under a 5-fold cross-validation algorithm. The hyper-parameters adjusted were learning rate (0.01, 0.04, 0.07, 0.1), number of trees (50, 100, 250, 500), and depth (1, 2, 3, 4). For each model we chose the best parameter combination based on the median precision obtained from the 5-fold cross validation, and trained the model within the train set using these parameters. The reliability of the model was validated based on the comparison between predictions made by the model within the 5-fold cross validation step, by comparing the predictive measures observed in the training model with the ones in the validation set. Then, we explored the differences in the prediction power between the training set after the 5-fold cross validation and the test set.

### Replication and cross-cohort validation strategy

To ensure the reproducibility of the model and to evaluate its reliability to classify diabetic patients, we performed a cross-cohort validation (**Supplemental Fig. 7**). To that end, we trained XGBoost in the UK Biobank cohort ^8^ and in 70KforT2D ^9^. Then, we used the trained models as predictors in the complementary cohort. We used weighted precision, recall and area under receiver operator curve (AUROC) to evaluate the reliability of the predictions. To further ensure the replicability of the model we used the UK Biobank trained model to predict in the GCAT|Genomes for life cohort ^44^ which consisted of the genotype of 2,627 variants in 4,988 individuals (450 diabetic, 4,538 non-diabetic), and assessed the trustworthiness of the model using the same reliability measures.

After ensuring reliability and replicability of the model, for the predicting variants suggested by the model in UKBiobank and 70KforT2D, we analyzed the overlap with GWAS significant signals from each cohort (**Supplemental Fig. 8**).

### Interpretation of ML results

The interpretation of the results was obtained by combining the internal tools provided by the interpretable model ^29^ and other external eXplainable Artificial Intelligence (XAI) tools ^36^.

First, since XGBoost is based on extreme gradient boosting trees, the results obtained were composed of a list of the most relevant variants for the method to do the classification with their corresponding scores, and the complete set of final decision trees including the decisions. After the test step, the method provides a list with the predictions and real observed values. The list of variants can be scored using two different measures, the weight, which is related to the number of times that the variant has been used to make a decision, or the gain, which corresponds to the accuracy value after adding the variant to the final model (**Fig. 3B**). The trees obtained represent combinations of variants, where the leaves are the variants in each group, and the branches are the decisions made by the method. The analysis of the complete set of decisions made during the training corresponds to finding differences between the variants genotype, thus responding to questions such as the variant being reference homozygous or alternate homozygous for a particular individual.

Then, to understand the relevance and the contribution of each variant in the prediction of disease or control groups at the individual patient level, we computed SHAP values over the test set predictions of each subgroup model ^36^. In brief, the SHAP algorithm estimates a value that represents the importance of a given feature in the model, for a given observation. This constitutes an XAI approach which improves the interpretability of any ML model. To further enhance the interpretation of the model results, we computed the median for each variant from the set of all its SHAPley absolute values on all individuals. Consequently, we were able to use this approach to prioritize variant relevance from the granularity of its contributions to disease. In addition, the directionality of individual SHAP values allowed us to infer if each genotype acted as a risk or protective factor for T2D onset (**Fig. 6A**).

We generated specific subgroup mixed-model predictors. To evaluate their performance in contrast with current PRS predictors, we calculated the genome-wide PRSs using T2D meta-analysis summary statistics resulting from our analysis in the UK Biobank. Multiple PRSs were generated using *p*-value thresholds ranging from 5×10^-8^ to 1.0 and using LD pruning parameters of 0.97 R^2^ over 250 kb windows. To evaluate the results we obtained multiple reliability measures including the confusion matrix and the weighted measures for AUROC, precision, recall and accuracy (**Supplemental Table 6**). Then, we used the general T2D mixed-model to classify diabetic individuals in T2D subgroups and compared the results using the same reliability measures. We additionally generated a combined predictor using the outcomes from T2D mixed-model and the subgroup-specific classifiers. This predictor relies on the sum of the predictions made by the mixed-models. More specifically, for each classifier *i* each individual is classified as diabetic with a probability 𝑝_𝑖,1_ and thus, it has a probability of 𝑝 = 1 − 𝑝_𝑖,0_ to be a control. Based on the combination of these probabilities, a score for being case or control can be assigned to each individual *j* so that, 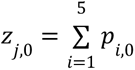 and 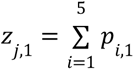. Then, if the score for an individual is above the average score for all individuals in any of 𝑧_𝑗,0_ or 𝑧_𝑗,1_, the individual can be assigned two predictions 𝑝𝑟𝑒𝑑_𝑗,0_ and 𝑝𝑟𝑒𝑑_𝑗,1_. If both 𝑝𝑟𝑒𝑑_𝑗,0_ and 𝑝𝑟𝑒𝑑_𝑗,1_ are 0, then, the individual is classified as control. If both 𝑝𝑟𝑒𝑑_𝑗,0_ and 𝑝𝑟𝑒𝑑_𝑗,1_ are 1, then, the individual is classified as case. Otherwise, it is considered that the individual cannot be classified with this methodology. This combined predictor was evaluated in 70KforT2D and compared with current T2D PRS.

## Data Availability

All data produced in the present work are contained in the manuscript and will be available online at zenodo

## Contributions

**Conceived the work:** David Torrents

**Designed the work:** David Torrents, Ignasi Morán, Juan Ramón González and Lorena Alonso-Parrilla

**Supervised the project:** David Torrents, Ignasi Morán and Lorena Alonso-Parrilla **Acquired the data:** Miguel Ángel Pérez-Elena, Cecilia Salvoro, Xavier Farré, Aikaterini Lymperidou, Natalia Blay and Lorena Alonso-Parrilla

**Prepared the data:** Lorena Alonso-Parrilla, Miguel Ángel Pérez-Elena, Cecilia Salvoro, Xavier Farré and Natalia Blay

**Analysed the data:** Lorena Alonso-Parrilla, Miguel Ángel Pérez-Elena, Lydia García, Mohammed Yousef Salem, Nicolás Gaitán, Leila Satari, Rodrigo Martín, Maedeh Mashhadikhan, Anthony Piron

**Interpreted the findings:** Lorena Alonso-Parrilla

**Supervised the findings:** Lorena Alonso-Parrilla, Ignasi Morán, Josep Lluis Berral, Rafael de Cid, Miriam Cnop and David Torrents

**Wrote the paper:** Lorena Alonso-Parrilla and David Torrents

**Revised the paper:** All authors

## Data availability

The complete summary statistics from this study have been deposited and are available to download at Zenodo (https://zenodo.org/uploads/15706418).

## Supplementary information

A supplementary information document is attached alongside this document.

## Acknowledgments

We thank the participants and investigators of the UK Biobank study who made this work possible (Resource Application Numbers 101646 and 83957). This work, integrated into the framework of PERTE for Vanguard Health, has been co-financed by the Spanish Ministry of Science, Innovation and Universities with funds from the European Union NextGenerationEU, from the Recovery, Transformation and Resilience Plan (PRTR-C17.I1) and from the Autonomous Community of Catalonia within the framework of the Biotechnology Plan Applied to Health, the Departament de Recerca i Universitats de la Generalitat de Catalunya (code: 2021 SGR 01626; 2021 SGR 01537), and the Science and Innovation Spanish Ministry under project PrevDis (PID2023-152867NB-I00). This work has been partially financed by the European Commission (EU-HORIZON NEARDATA GA.101092644). This study makes use of data generated by the GCAT |Genomes for Life Project. Cohort study of the Genomes of Catalonia. GCAT linked health registers were provided by the Catalan Agency for Quality and Health Assessment (PADRIS Program). IGTP is part of the CERCA Program / Generalitat de Catalunya. A full list of the investigators who contributed to the generation of the data is available from www.genomesforlife.com. This project has received funding from the Generalitat de Catalunya through the CERCA Program and the Consolidated Group on HEALTH ANALYTICS (2021 SGR 01563). We also acknowledge support from the grant CEX2023-0001290-S funded by MCIN/AEI/ 10.13039/501100011033, and support from the Generalitat de Catalunya through the CERCA Program. L.A-P fellowship within the “Generación D” initiative, Red.es, Ministerio para la Transformación Digital y de la Función Pública, for talent attraction (C005/24-ED CV1). Funded by the European Union NextGenerationEU funds, through PRTR. N.G fellowship under the FPU23/03413 (Formación de Profesorado Universitario) scholarship by the Spanish Ministry of Science, Innovation and Universities. X.F is funded by TED2021-130626B-I00. A.L is funded by the Ministry of Science and Innovation with funding from the European Union NextGenerationEU, the Recovery, Transformation and Resilience Plan (PRTR-C17.I1), and the Autonomous Community of Catalonia. M.C. is funded by Walloon Region strategic axis Fonds de la Recherche Scientifique (FRFS)–Walloon Excellence in Life Sciences and Biotechnology (WELBIO). The technical support group from the Barcelona Supercomputing Center is gratefully acknowledged. Finally, we thank all the Computational Genomics group at the BSC for their helpful discussions and valuable comments on the manuscript.

## Notes

### Competing Interest Statement

Cecilia Salvoro was employed by AstraZeneca plc, until July 2025, and by Genomics Ltd. later on

### Author Declarations

FUSION (dbGaP phs000867.v1.p1) GENEVA (dbGaP phs000091.v2.p1) NUgene (dbGaP phs000237.v1.p1) GERA (dbGaP phs000788.v2.p3) WTCCC (https://www.wtccc.org.uk/) UK Biobank (https://www.ukbiobank.ac.uk/) GCAT (http://www.gcatbiobank.org/investigadors/en_gcat-data-access/)

## References

1. Zhou, B. et al. Worldwide trends in diabetes prevalence and treatment from 1990 to 2022: a pooled analysis of 1108 population-representative studies with 141 million participants. The Lancet 404, 2077–2093 (2024).

2. IDF Diabetes Atlas 2025. Diabetes Atlas https://diabetesatlas.org/resources/idf-diabetes-atlas-2025/.

3. Ahlqvist, E. et al. Novel subgroups of adult-onset diabetes and their association with outcomes: a data-driven cluster analysis of six variables. Lancet Diabetes Endocrinol. 6, 361–369 (2018).

4. Florez, J. C. Advancing precision medicine in type 2 diabetes. Lancet Diabetes Endocrinol. 12, 87–88 (2024).

5. Udler, M. S. et al. Type 2 diabetes genetic loci informed by multi-trait associations point to disease mechanisms and subtypes: A soft clustering analysis. PLoS Med. 15, (2018).

6. Mansour Aly, D., et al. Genome-wide association analyses highlight etiological differences underlying newly defined subtypes of diabetes. Nat. Genet. 53, 1534–1542 (2021).

7. Misra, S. et al. Precision subclassification of type 2 diabetes: a systematic review. Commun. Med. 3, 1–19 (2023).

8. Sudlow, C. et al. UK Biobank: An Open Access Resource for Identifying the Causes of a Wide Range of Complex Diseases of Middle and Old Age. PLoS Med. 12, 1001779 (2015).

9. Bonàs-Guarch, S. et al. Re-analysis of public genetic data reveals a rare X-chromosomal variant associated with type 2 diabetes. Nat. Commun. 9, 321 (2018).

10. Guindo-Martínez, M., Amela, R., &, et al. The impact of non-additive genetic associations on age-related complex diseases. Nat. Commun. 12, 2436 (2021).

11. Alonso, L., Piron, A., Morán, I., &, et al. TIGER: The gene expression regulatory variation landscape of human pancreatic islets. Cell Rep. 37, 109807 (2021).

12. Boomsma, D. I. et al. The Genome of the Netherlands: Design, and project goals. Eur. J. Hum. Genet. 22, 221–227 (2014).

13. The 1000 Genomes Project Consortium. A global reference for human genetic variation. Nature 526, 68–74 (2015).

14. The Haplotype Reference Consortium. A reference panel of 64,976 haplotypes for genotype imputation. Nat. Genet. 48, 1279–1283 (2016).

15. The UK10K Consortium. The UK10K project identifies rare variants in health and disease. Nature 526, 82–90 (2015).

16. Valls-Margarit, J., Galván-Femenía, I., Matias, D. & Al., E. GCAT|Panel, a comprehensive structural variant haplotype map of the Iberian population from high-coverage whole-genome sequencing. Nucleic Acids Res. 1, 13–14 (2022).

17. Slieker, R. et al. Replication and cross-validation of type 2 diabetes subtypes based on clinical variables: an IMI-RHAPSODY study. Diabetologia 64, 1982–1989 (2021).

18. Xie, J. et al. Validation of type 2 diabetes subgroups by simple clinical parameters: a retrospective cohort study of NHANES data from 1999 to 2014. BMJ Open 12, e055647 (2022).

19. Kautzky-Willer, A., Leutner, M. & Harreiter, J. Sex differences in type 2 diabetes. Diabetologia 66, 986–1002 (2023).

20. Mbatchou, J. et al. Computationally efficient whole-genome regression for quantitative and binary traits. Nat. Genet. 2021 537 53, 1097–1103 (2021).

21. Magi, R., Lindgren, C. M. & Morris, A. P. Meta-analysis of sex-specific genome-wide association studies. Genet. Epidemiol. 34, 846–853 (2010).

22. Mägi, R. & Morris, A. P. GWAMA: Software for genome-wide association meta-analysis. BMC Bioinformatics 11, (2010).

23. Mahajan, A. et al. Multi-ancestry genetic study of type 2 diabetes highlights the power of diverse populations for discovery and translation. Nat. Genet. 54, 560–572 (2022).

24. Sollis, E. et al. The NHGRI-EBI GWAS Catalog: knowledgebase and deposition resource. Nucleic Acids Res. 51, D977–D985 (2023).

25. Ahlqvist, E., Prasad, R. B. & Groop, L. Subtypes of Type 2 Diabetes Determined From Clinical Parameters. Diabetes 69, 2086–2093 (2020).

26. Mahajan, A. et al. Fine-mapping type 2 diabetes loci to single-variant resolution using high-density imputation and islet-specific epigenome maps. Nat. Genet. 50, 1505–1513 (2018).

27. Li, M. et al. Trends in insulin resistance: insights into mechanisms and therapeutic strategy. Signal Transduct. Target. Ther. 7, 1–25 (2022).

28. Yüzbaşıoğulları, A. B. et al. Sex-specific associations of TCF7L2 variants with fasting glucose, type 2 diabetes and coronary heart disease among Turkish adults. Anatol. J. Cardiol. 24, 326–333 (2020).

29. Chen, T. & Guestrin, C. XGBoost: A Scalable Tree Boosting System. in *Proceedings of the 22nd ACM SIGKDD International Conference on Knowledge Discovery and Data Mining* 785–794 (Association for Computing Machinery, New York, NY, USA, 2016). doi:10.1145/2939672.2939785.

30. Matsuzaka, T. & Shimano, H. Elovl6: a new player in fatty acid metabolism and insulin sensitivity. J. Mol. Med. 87, 379–384 (2009).

31. Zhao, H. et al. Elovl6 Deficiency Improves Glycemic Control in Diabetic db/db Mice by Expanding β-Cell Mass and Increasing Insulin Secretory Capacity | Diabetes | American Diabetes Association. Diabetes 66, 1833–1846 (2017).

32. Pasquali, L. et al. Pancreatic islet enhancer clusters enriched in type 2 diabetes risk-associated variants. Nat. Genet. 46, 136–143 (2014).

33. Miguel-Escalada, I. et al. Human pancreatic islet three-dimensional chromatin architecture provides insights into the genetics of type 2 diabetes. Nat. Genet. 51, 1137–1148 (2019).

34. The GTEx Consortium. The GTEx Consortium atlas of genetic regulatory effects across human tissues. Science 369, 1318–1330 (2020).

35. 60th EASD Annual Meeting of the European Association for the Study of Diabetes. Diabetologia 67, 1–593 (2024).

36. Lundberg, S. M. & Lee, S.-I. A unified approach to interpreting model predictions. in Proceedings of the 31st International Conference on Neural Information Processing Systems 4768–4777 (Curran Associates Inc., Red Hook, NY, USA, 2017).

37. Marselli, L. et al. Persistent or Transient Human β Cell Dysfunction Induced by Metabolic Stress: Specific Signatures and Shared Gene Expression with Type 2 Diabetes. Cell Rep. 33, 108466 (2020).

38. García-Pérez, R. et al. The landscape of expression and alternative splicing variation across human traits. Cell Genomics 3, 100244 (2023).

39. Burton, P. R. et al. Genome-wide association study of 14,000 cases of seven common diseases and 3,000 shared controls. Nature 447, 661–678 (2007).

40. Colditz, G. A. & Hankinson, S. E. The nurses’ health study: Lifestyle and health among women. Nat. Rev. Cancer 5, 388–396 (2005).

41. Ghosh, S. et al. The Finland-United States investigation of non-insulin-dependent diabetes mellitus genetics (FUSION) study. I. An autosomal genome scan for genes that predispose to type 2 diabetes. Am. J. Hum. Genet. 67, 1174–1185 (2000).

42. Gottesman, O. et al. The Electronic Medical Records and Genomics (eMERGE) Network: Past, present, and future. Genet. Med. 15, 761–771 (2013).

43. Kvale, M. N. et al. Genotyping informatics and quality control for 100,000 subjects in the genetic epidemiology research on adult health and aging (GERA) cohort. Genetics 200, 1051–1060 (2015).

44. Obón-Santacana, M. et al. GCAT|Genomes for life: a prospective cohort study of the genomes of Catalonia. BMJ Open 8, 18324 (2018).

45. Purcell, S. et al. PLINK: A tool set for whole-genome association and population-based linkage analyses. Am. J. Hum. Genet. 81, 559–575 (2007).

46. Bycroft, C. et al. The UK Biobank resource with deep phenotyping and genomic data. Nat. 2018 5627726 562, 203–209 (2018).

47. Delaneau, O., Zagury, J.-F., Robinson, M. R., Marchini, J. L. & Dermitzakis, E. T. Accurate, scalable and integrative haplotype estimation. Nat. Commun. 10, 5436 (2019).

48. Howie, B. N., Donnelly, P. & Marchini, J. A Flexible and Accurate Genotype Imputation Method for the Next Generation of Genome-Wide Association Studies. PLOS Genet. 5, e1000529 (2009).

49. Dennis, J. M., Shields, B. M., Henley, W. E., Jones, A. G. & Hattersley, A. T. Disease progression and treatment response in data-driven subgroups of type 2 diabetes compared with models based on simple clinical features: an analysis using clinical trial data. Lancet Diabetes Endocrinol. 7, 442–451 (2019).

50. Kahkoska, A. R. et al. Validation of distinct type 2 diabetes clusters and their association with diabetes complications in the DEVOTE, LEADER and SUSTAIN-6 cardiovascular outcomes trials. Diabetes Obes. Metab. 22, 1537–1547 (2020).

51. Katki, H. A. et al. Increase in power by obtaining 10 or more controls per case when type-1 error is small in large-scale association studies. BMC Med. Res. Methodol. 23, 153 (2023).

52. Gauderman, W. J. Sample size requirements for matched case-control studies of gene-environment interaction. Stat. Med. 21, 35–50 (2002).

53. McLaren, W. et al. The Ensembl Variant Effect Predictor. Genome Biol. 17, 122 (2016).

54. Dyer, S. C. et al. Ensembl 2025. Nucleic Acids Res. 53, D948–D957 (2025).

55. Raudvere, U. et al. g:Profiler: a web server for functional enrichment analysis and conversions of gene lists (2019 update). Nucleic Acids Res. 47, W191–W198 (2019).

56. Sherman, B. T. et al. DAVID: a web server for functional enrichment analysis and functional annotation of gene lists (2021 update). Nucleic Acids Res. 50, W216–W221 (2022).

57. Zabaneh, D. et al. Fine mapping genetic associations between the HLA region and extremely high intelligence. Sci. Rep. 7, 41182 (2017).

58. Piron, A. et al. Identification of novel type 1 and type 2 diabetes genes by co-localization of human islet eQTL and GWAS variants with colocRedRibbon. 2024.10.19.24315808 Preprint at 10.1101/2024.10.19.24315808 (2024).

59. Pedregosa, F. et al. Scikit-learn: Machine Learning in Python. J Mach Learn Res 12, 2825–2830 (2011).

